# Precise modeling of task-related sensorimotor activation based on simultaneous surface electromyography

**DOI:** 10.64898/2026.02.01.26345279

**Authors:** Monika Jasenská, Pavel Hok, Martin Kojan, Ondřej Burkot, Barbora Kolářová, Aleš Holobar, Petr Hluštík

## Abstract

**Objective:** To evaluate central and peripheral correlates of motor control using functional magnetic resonance imaging (fMRI) and surface electromyography (EMG), with a focus on the added value of EMG-informed analysis during movement of the lower limb in healthy controls.

**Methods:** Twenty participants performed dorsi-/plantarflexion of the ankle (Ankle) and gait imagery (GI) in a block design during fMRI. Accelerometry (Acc) and surface EMG from tibialis anterior (TA) activity were recorded and included as regressors in five analysis models, either with or without temporal derivative (TD) to account for time shift in the task regressor. Voxel-wise analyses complemented by post-hoc region-of-interest (ROI) analyses were performed to compare the amount of variability explained by the models.

**Results:** Inclusion of either Acc or EMG on top of the task regressor explained robustly fMRI signal variability in the primary sensorimotor cortices. On top of Acc, EMG additionally explained activation variability mainly in the contralateral thalamus and the secondary somatosensory cortex (S2). This effect was, however, mainly driven by spontaneous signal fluctuations at rest and during imagery. Comparisons between models with and without TD revealed consistent differences in the cerebellum and thalamus across tested models, suggesting that subcortical structures may involve transient signal changes when switching between movement and rest.

**Conclusion:** Including EMG in fMRI analysis enhances specificity in detecting motor-related brain activity and enables differentiation of spontaneous or unpredicted motor behavior. TD improved signal detection in the primary sensorimotor cortices, but may have a detrimental effect on signal detection in other, mostly subcortical regions, likely reflecting their different temporal signal dynamics.

## 1. Introduction

Objective functional magnetic resonance imaging (fMRI) biomarkers for gait impairment or motor deficits in general are continuing to be an unmet goal of neuroimaging research (Bullmore and Sporns, 2012; Rocca et al., 2022). Despite its advantages, fMRI suffers from a lack of technical standardization, resulting in limited use of fMRI biomarkers in clinical routine (Soares et al., 2016). Insufficient control of task performance is another common pitfall often hindering the interpretability in task-related fMRI (Bullmore and Sporns, 2012).

Motor task-related EMG-fMRI has emerged as a promising approach to overcome these limitations. By combining the spatial sensitivity of fMRI with the temporal specificity and muscle-level resolution of surface electromyography (EMG), it offers the potential to detect abnormalities associated even with subtle gait disturbances. EMG records electrical muscle activity and among electrophysiological measures correlates most closely with overt motion (Kranjec and Holobar, 2019; Kramberger and Holobar, 2021). Even during movement imagery, EMG can capture changes reflecting facilitation or inhibition of relevant muscles (Haltmar et al., 2025; Kolářová et al., 2016). While inertial measurement units (IMUs) can provide objective verification of movement execution through kinematic data, they do not capture underlying neuromuscular strategies. In contrast, EMG delivers complementary information about muscle engagement and motor control processes that are not evident from basic movement verification alone (Jasenská et al., 2025). EMG may thus reveal relationships between brain activation and peripheral muscle engagement not only due to voluntary motion, but also reflecting the modulatory influences of higher order motor centers.

A promising basis for such development can be found in several fMRI studies that have examined the functional anatomy of motor control during ankle movement, a critical component of the gait cycle (Dobkin et al., 2004; MacIntosh et al., 2004). Specifically, repetitive ankle dorsiflexion has proven to be an effective paradigm to assess supraspinal sensorimotor networks involved in the control of walking (Dobkin et al., 2004). Given its simplicity and reproducibility, this task is particularly well-suited for EMG-fMRI applications, allowing for targeted EMG measurements of the tibialis anterior (TA), the primary muscle contributing to dorsiflexion (Francis et al., 2009; MacIntosh et al., 2007), which plays key roles in gait.

Studies using combined EMG-fMRI have shown that EMG-informed fMRI models are more accurate with respect to movement onset or by controlling for motor output and therefore more sensitive to activation (Francis et al., 2009; MacIntosh et al., 2007; Shitara et al., 2013), and that the variation in EMG explains variation in areas exerting modulatory control over movements (Haruno et al.,2012; van Duinen et al., 2008; van Rootselaar et al., 2007) or activation associated with involuntary manifestations of movement disorders (Daly et al., 2008; Richardson et al., 2006; van Rootselaar et al., 2007). However, fMRI model designs using EMG differ considerably among studies: the EMG signal can be included in the general linear model (GLM) as a raw time-series (van Duinen et al., 2008) or more often convolved with hemodynamic response function, or additionally orthogonalized with respect to the task regressor to obtain a so-called residual EMG (rEMG) reflecting motor output variation during task performance (Broersma et al., 2016; van Rootselaar et al., 2007, 2008). The choice of optimal EMG-fMRI model that would maximize the explained variance in the blood oxygenation level-dependent (BOLD) signal remains thus unclear, especially with respect to the lower limb EMG as most previous studies were conducted in the upper limbs ([Daly et al., 2008; Richardson et al., 2006; Sehm et al., 2010; Sharifi et al., 2022; van Duinen et al., 2008; van Rootselaar et al., 2007, 2008] vs. [Francis et al., 2009; MacIntosh et al., 2007]). Moreover, while IMUs provide useful kinematic information, particularly through their acceleration (Acc) signals, the utility of adding simultaneous EMG on top of IMU-derived acceleration has not been rigorously evaluated.

While previous studies have demonstrated that orthogonalized rEMG can identify movement-related brain activity independently of task timing (van Rootselaar et al., 2007, 2008), no systematic comparison has been made between rEMG and models that do not use EMG. Furthermore, prior research has focused almost exclusively on motor execution, leaving the potential contribution of EMG variability during rest or motor imagery unaddressed. Therefore, we aimed to systematically compare different EMG-fMRI modeling strategies, including a reference model without EMG regressors, models using orthogonalized EMG signals (rEMG), and models with orthogonalized IMU signal with and without rEMG. We further aimed to extend the application of rEMG by evaluating its role not only during repetitive active motor tasks but also in a more complex paradigm involving different rest and imagery conditions. In such designs, controlling for spontaneous fluctuations during rest or imagery by separate modeling of active and resting states may assess whether EMG fluctuations reflect meaningful neural activity beyond overt movement (possibly engaging distinct neural populations), and whether rEMG from non-task periods carries unique explanatory power in modeling BOLD responses. We hypothesized that including EMG information would improve the explained variance of the BOLD signal, particularly for modeling movement-related brain activity in the lower limb. Specifically, we were interested (h1) whether Acc or EMG (either as a whole or separately modeling the active task) explain any BOLD signal variability; (h2) explained variability differs between models with Acc and EMG; (h3) EMG explains additional variability on top of Acc; (h4) EMG during rest explains any variability in the model.

One of the possible mechanisms, by which EMG improves overall model sensitivity, is the reduction of residuals in the case of slightly time-shifted hemodynamic response function (HRF) due to delayed or premature movement onset. For similar reasons, temporal derivatives (TD) of the task regressors have been often incorporated in the GLM (Calhoun et al., 2004). TD may correct for slight timing mismatches and variable hemodynamic delays. Simulation results further confirm that including the TD improves model fit and parameter estimation, especially under conditions of temporal variability (Pernet, 2014). While the motivation to include TD overlaps with the rationale for inclusion of rEMG, the combined use of rEMG regressors and TD has not been properly evaluated. Hence, we additionally aimed to investigate the impact of temporal modeling by systematically comparing models using EMG regressors with and without TD. We hypothesized that (h5) including TD would lead to better model performance by improving the sensitivity and accuracy of EMG-related BOLD signal detection.

## 2. Materials and methods

### 2.1 Participants

Twenty-three healthy subjects (32.5±7.5 years; 10 women, 13 men) underwent a single 3T MRI examination with concomitant acquisition of EMG and kinematic data. Exclusion criteria were: any motor disability, musculoskeletal or neurological impairments affecting the lower limbs, and contraindications for MRI (e.g., non-compatible implants, claustrophobia, or other safety-related issues).

The study was carried out in accordance with the World Medical Association Declaration of Helsinki. Written informed consent was obtained from all participants prior to their inclusion in the study and study protocol was approved by the local ethics committee (Ethics Committee of the University Hospital and the Faculty of Medicine and Dentistry of Palacký University Olomouc, Czech Republic) approval number 48/23.

### 2.2 Experimental task and setting (simultaneous EMG-fMRI)

The task-based fMRI protocol consisted of two equal 10-min paradigms. Here, only the first 10-min repetition is further reported. The paradigm involved 4 different conditions, each repeated 5 times in a block design (Fig. 1): (1) 9 cycles of tone-paced dorsiflexion alternating with plantarflexion of the ankle (Ankle) using non-dominant lower limb; (2) gait imagery (GI); (3) rest without tones (silence); and (4) rest with tones (beeps). Each block lasted 30 s, including a 3-s audiovisual instruction (inst) to initiate each 27-s task.

**Fig. 1.**
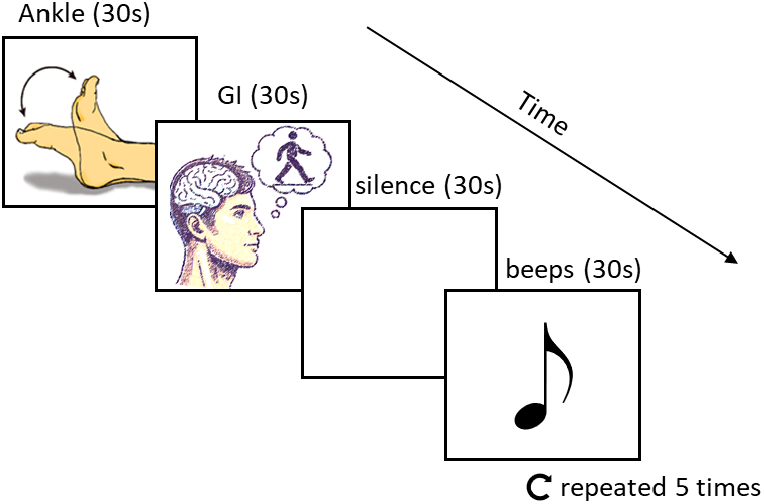
fMRI block paradigm. Active and passive tasks for selective activation of motor areas; GI – gait imagery.

Both active tasks were briefly trained outside the scanner room. For GI training, they alternated between actual walking and imagining walking while seated for 30 s intervals over 2 cycles. Instructions focused on motor and kinesthetic aspects of imagery rather than visual, emphasizing actual gait rather than optimal gait (Boyne et al., 2020). Although GI was reflected in the analysis (see below), this report further focuses exclusively on activation during Ankle condition.

Inside the scanner bore, participants lay supine with their knees flexed at 120-140°. Subjects were instructed to perform the ankle dorsi-/plantarflexion with their non-dominant lower limb. The actual ankle movement range was measured with a goniometer in the scanner before the fMRI experiment. Each subject’s head was stabilized with adjustable padded restraints on both sides, along with additional restraints at the shoulders and hips. Participants were instructed to remain as still as possible throughout the experiment. The lights remained on throughout the acquisition and instructions were delivered via headphones. Subjects were continuously monitored and recordings of physiological signals were observed during the scanning to assess compliance.

#### 2.2.1 EMG and physiology data acquisition

Physiological signals, namely surface electromyography, heart rate, respiration rate and kinematic data (accelerometry - Acc) were acquired simultaneously (Brain Products GmbH, Gilching, Germany) during BOLD scanning and synchronized using markers from the scanner’s gradient system, enabling temporal alignment with fMRI volumes. This setup allows for offline removal of physiological artifacts and supports the analysis of autonomic responses and muscle activation.

During fMRI acquisition, the EMG signal was sampled at 5000 Hz/channel using a digital recording MR-compatible system (BrainAmp ExG MR, Brain Products, Gilching, Germany). The EMG electrode pairs (Arbo™ H135TSG, Kendall, Mansfield, USA) were placed on the non-dominant lower limb, specifically over the TA muscle, in a bipolar recording configuration. The electrode placement was performed following the SENIAM guidelines (Hermens et al., 2000). The sensors were attached to the skin using double-sided adhesive tape. The electrode wires were twisted per electrode pair to minimize the differential effect of the magnetic field on the EMG leads.

Triaxial acceleration data were obtained from the dorsal side of the foot in the direction of the metatarsus. Heart rate (ECG, single-lead) and respiration (respiratory belt) were monitored during BOLD scanning (BrainAmp ExG MR, Brain Products, Gilching, Germany).

#### 2.2.2 fMRI data acquisition

fMRI data were acquired using a 3T scanner (Siemens Prisma, Erlangen, Germany) with a standard 64-channel head and neck coil in the Multimodal and Functional Imaging Laboratory (MAFIL), Central European Institute of Technology (CEITEC) in Brno. The MRI protocol included a task-related blood oxygenation level-dependent (BOLD) fMRI data acquisition using a T_2_*-weighted EPI sequence [48 slices, repetition time (TR) = 695 ms, echo time (TE) = 30 ms, 72×72 matrix, field of view (FOV) = 216×216, slice thickness 3 mm, voxel size 3×3×3 mm, axial slice orientation, multi-band (MB) = 4, parallel acquisition technique (PAT) = 2, 880 volumes resulting in time of acquisition of 10 min 26 s (including saturation scans). A T_1_-weighted anatomical scan (208 slices, TR = 2300 ms, TE = 3.68 ms, scanning matrix 256×256, FOV = 230×230, slice thickness 0.9 mm, axial slice orientation) along with fieldmaps (two magnitude components, one phase component, 48 slices, TR = 622 ms, TE 1 = 4.92 ms, TE 2 = 7.38 ms, 72×72 matrix, FOV = 216×216, slice thickness 3 mm, voxel size 3×3×3 mm, axial slice orientation) were also acquired.

#### 2.2.3 EMG and physiology data pre-processing

The entire analysis pipeline is schematically presented in Fig. 2. In detail, we preprocessed the physiology data using BrainVision Analyzer 2.2 (Brain Products, Gilching, Germany) following the same pipeline for all subjects. Scanner-related gradient artifacts were removed using the Sliding Average Calculation method (Allen et al., 2000), with continuous correction applied at each repetition time (TR = 695 ms). Correction templates were computed from a sliding average of 173 intervals, baseline correction was enabled and all channels were included. After artifact removal, the data were down sampled by a factor of 5 to 1000 Hz, and a total of 880 scanner artifacts were detected and corrected. All physiological data were stored for further processing and analysis in MATLAB R2024b (Mathworks Inc., Natick, USA). Preprocessing also included identification of the MRI scan onset using synchronization markers, as well as identification of the beginning of physiological signal recordings.

**Fig. 2.**
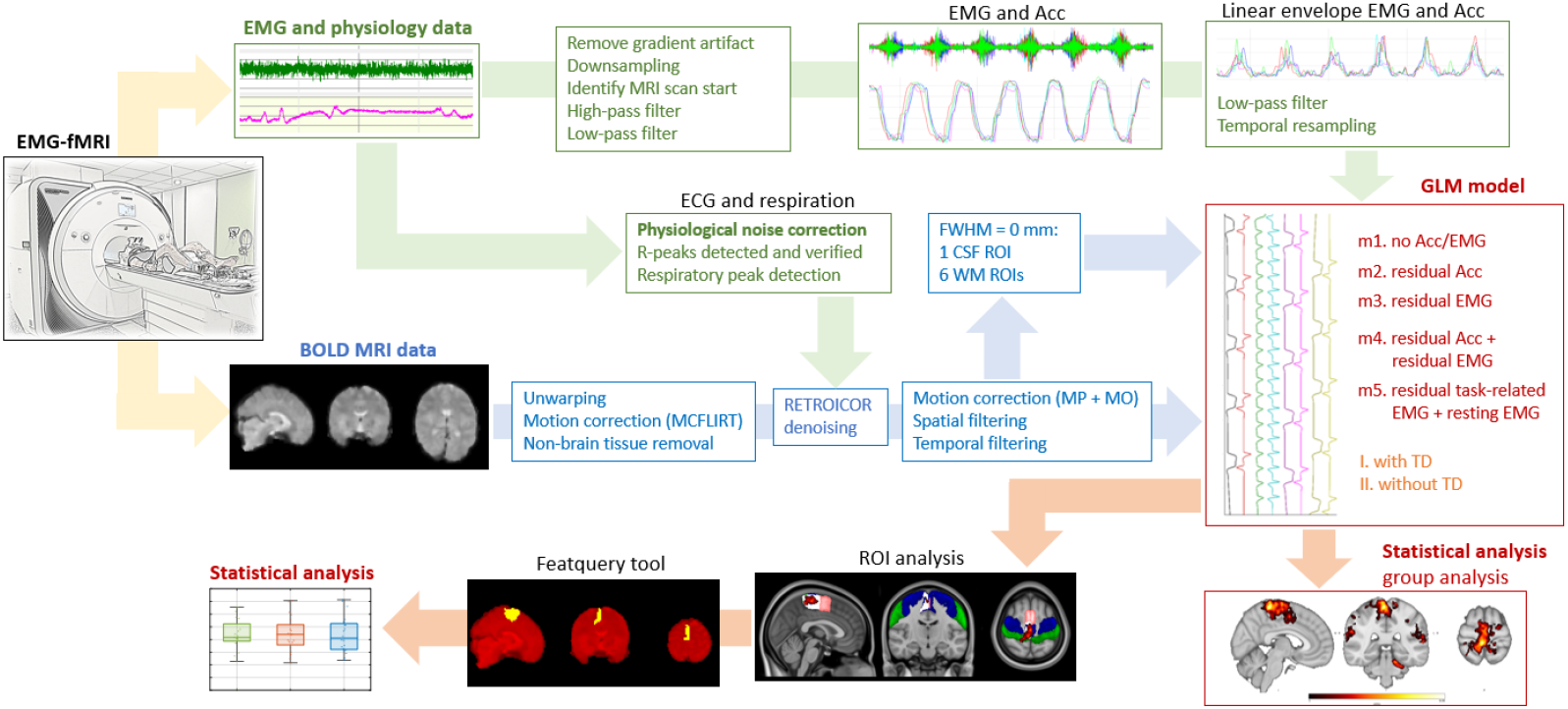
Block diagram of the data preprocessing and analysis pipeline. Abbreviation: Acc – accelerometry; BOLD – blood-oxygen-level–dependent; CSF – cerebrospinal fluid; EMG – electromyography; ECG – electrocardiography, fMRI – functional magnetic resonance imaging; FWHM – full width at half maximum; GLM – general linear model; MO – motion outliers; MP – motion parameters; MRI – magnetic resonance imaging; RETROICOR – RETROspective Image-based CORrection; ROI – region of interest; TA – tibialis anterior; TD – temporal derivative; WM – white matter.

EMG data were preprocessed according to the Consensus for Experimental Design in Electromyography (CEDE) guidelines (Dick et al., 2024; Besomi et al., 2020). We filtered it using two zero-phase 4th-order Butterworth filters, a 20 Hz high-pass filter, and a 250 Hz low-pass filter (Merletti and Cerone, 2020; De Luca et al., 2010). The signals were then full-wave rectified and, afterwards, low-pass filtered at 2 Hz to create an amplitude envelope of the EMG signal (Clancy et al., 2023; Francis et al., 2009). The calculated EMG amplitude envelopes were resampled to match the temporal resolution of fMRI time series and used as regressors in the GLM for fMRI analysis (van Rootselaar et al., 2007).

The accelerometric data reflected the participants’ motor performance during the fMRI acquisition. Triaxial accelerometry data were filtered using the 4th-order Butterworth high-pass filter with a cutoff frequency of 0.3 Hz and a low-pass filter with a cutoff frequency of 10 Hz (Budini et al., 2018). The input data were processed by combining the signals from all three axes (acceleration vector) into a Euclidean norm (Voisard et al., 2024), representing the total acceleration magnitude across all directions in 3D space. The signals were then full-wave rectified and low-pass filtered at 2 Hz to create the Acc amplitude envelope. The Acc amplitude envelopes were resampled to match the temporal resolution of fMRI time series and used as regressors in the GLM for fMRI analysis.

The ECG recordings and the respiratory curve were processed for physiological noise correction, with R-peaks detected and verified in the ECG, maximum peaks identified in the respiratory curve. These detected features were then utilized and prepared for further processing according to the RETROICOR denoising procedure (RETROspective Image-based CORrection; Glover et al., 2000).

#### 2.2.4 BOLD MRI data pre-processing

The fMRI data were processed using FEAT version 6.00, part of FSL (FMRIB’s Software Library), version 6.0.5. (Jenkinson et al., 2012). Three subjects were removed from group analysis due to missing anatomical images, lack of relevant activations due to a subject falling asleep or noisy EMG recordings. The initial preprocessing consisted of correction of B0 distortions (BOLD MRI data unwarping using acquired field map images), motion correction and non-brain tissue removal using Brain Extraction Tool (BET), part of FSL (Smith, 2002). After these steps, nuisance signal regressors based on RETROICOR (Glover et al., 2000) were regressed out from the data. During pre-processing, an affine registration matrix between the functional images and the respective structural image was obtained using FLIRT (Jenkinson and Smith, 2001, Jenkinson et al., 2002, Greve et al., 2009) and a non-linear transformation between the structural space and the Montreal Neurological Institute (MNI) 152 standard space was calculated using FNIRT (Andersson et al., 2007), part of FSL. Next, high-pass temporal filtering with a sigma of 120.0 s and spatial filtering with an FWHM of 6 mm were applied. Finally, nuisance signal from cerebro-spinal fluid (CSF) and 6 regions of interest (ROIs) within the white matter (WM) were extracted from the pre-processed images from a parallel model without spatial filtering applied to provide regressors for GLM (Bartoň et. al., 2019).

### 2.3 Statistical analysis of BOLD imaging data

#### 2.3.1 First-level model

Voxel-wise GLM analysis was carried out using FILM (Woolrich et al., 2001). At the single-subject level, each model consisted of at least 4 main regressors to separately model activations evoked by ankle movement, gait imagery, rest with auditory pacing, and auditory instructions (for all blocks). Up to two additional regressors per model included preprocessed EMG and/or Acc recordings. An empty dummy EMG regressor was included to preserve model structure where applicable. In total, five parallel analysis models were created to assess the effect of inclusion of EMG/Acc recordings on resulting activation maps (Fig. 2). In the first model (m1, “standard” model), no physiological signal regressor was used. In the second model (m2, “rAcc” model), the Acc regressor orthogonalized with respect to the ankle movement regressor (residual Acc, or rAcc) was used (van Rootselaar et al., 2007). In the third model (m3, “rEMG” model), the EMG regressor orthogonalized with respect to the ankle movement regressor (rEMG) was used (van Rootselaar et al., 2007). In the fourth model (m4, “rAcc+orEMG” model), rAcc model was expanded by addition of rEMG, which was additionally orthogonalized with respect to the rAcc (orEMG). In the fifth model (m5, “split EMG” model), the EMG regressor was split into two components, namely resting-state EMG (rest EMG) without orthogonalization and orthogonalized task-related rEMG (trEMG), which consisted of the entire Ankle condition plus 3 seconds before and after the condition and was orthogonalized with respect to the ankle movement regressor. The epochs removed from each regressor were replaced by the mean of the resting-state epochs.

First-level contrasts were defined to evaluate the main effects of ankle movement (standard model m1), the independent contribution of the rAcc or rEMG regressors (models m2 and m3), rEMG orthogonalized with respect to rAcc (m4), and effects of trEMG and rest EMG (m5).

Since one of the reasons to include TD is to account for individual variability in task performance (movement onset delay), which is also reflected by EMG, a parallel set of models without TD of the first regressor (ankle movement) was also evaluated (models m1’-m5’). TD of the remaining regressors were always included to account for non-uniform slice timing and potential HRF delays.

#### 2.3.2 Second-level voxel-wise analysis

Group-level analysis was performed in FEAT using a mixed-effects model (FLAME 1+2), which accounts for both within- and between-subject variability among the models, providing group-mean activation maps and pairwise contrasts. Statistical maps were thresholded using a cluster-forming threshold of Z = 3.1, with cluster-wise correction at p < 0.05 (family-wise error corrected). To ensure that only brain tissue was included in the analysis, a standard whole-brain mask in MNI space was applied.

One-sample t-test contrasts were used to generate mean activation maps (testing hypotheses h1, h3, h4). Specifically, activation maps were computed for the main effects of rAcc and rEMG (hypothesis h1), main effect of orEMG (hypothesis h3), and main effects of trEMG and rest EMG (hypothesis h4). Paired t-test contrasts for comparisons among the models (testing hypotheses h2 and h5) were performed where appropriate. For hypothesis h2, main effects of rAcc, rEMG, and trEMG were compared with each other. For hypothesis h5, main effects of the ankle movement regressor, rAcc, rEMG, orEMG, trEMG and rest EMG from models m1-m5 were compared with the same regressor from corresponding models m1’-m5’.

### 2.4 Post-hoc region of interest analysis

In a post-hoc statistical analysis, mean Z-scores (representing the contrasts defined above) were extracted using Featquery tool (part of FSL) from cortical and subcortical regions (Fig. 3) defined as regions of interest (ROIs) according to the Brainnetome Atlas (https://atlas.brainnetome.org/), including the sensorimotor cortex (SMC; left hemisphere ROIs 65, 67 and right hemisphere 66, 68), supplementary motor area (SMA; ROIs 9 and 10), secondary somatosensory cortex (S2; left hemisphere ROI 145 and right hemisphere 146), and the thalamus (left hemisphere ROIs 233, 235, 245 and right hemisphere ROIs 234, 236, 246). Additionally, cerebellar regions were extracted from the MNI-based cerebellar atlas (cerebellum; ROI 2). These data were processed using MATLAB R2024b (MathWorks, Natick, MA, USA).

**Fig. 3.**
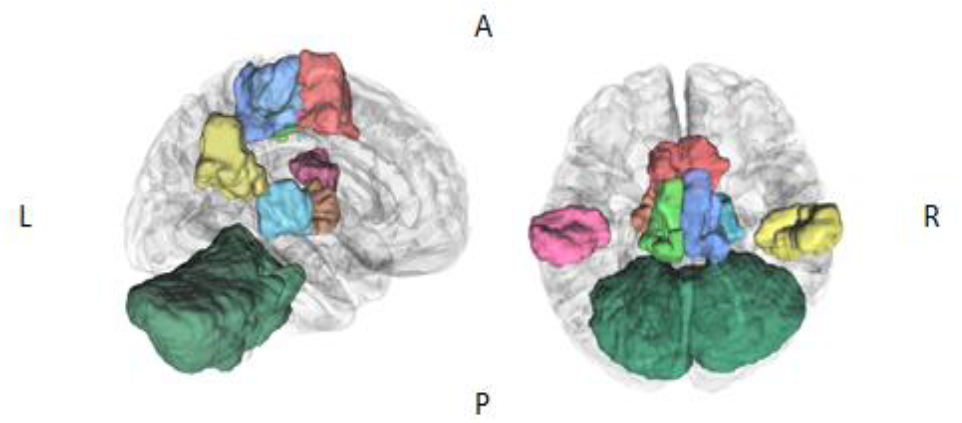
Cortical and subcortical regions defined as regions of interest (ROIs) according to the Brainnetome Atlas. Sensorimotor cortex (SMC; left hemisphere green and right hemisphere blue), supplementary motor area (SMA; red), secondary somatosensory cortex (S2; left hemisphere pink and right hemisphere yellow), the thalamus (left hemisphere orange and right hemisphere turquoise) and cerebellum (dark green).

Non-parametric Wilcoxon signed-rank tests with null hypothesis Z = 0 were used to test hypotheses h1, h3, and h4. Paired Wilcoxon signed-rank tests were performed to compare models of m2, m3, and m5 (hypothesis h2), or models m1-m5 with models m1’-m5’ (hypothesis h5).

## 3. Results

### 3.1 Behavioral data

Average maximum dorsiflexion was 37.7 ± 11.4° and average maximum plantarflexion was 23.6 ± 10.0°. During the Ankle condition, root mean square EMG activity in the TA muscle was 55.4 ± 8.1 μV, and it was associated with a median range of variation in foot accelerations of 0.9 g along the antero-posterior axis, as measured by the foot-mounted accelerometer. In total, 3 subjects were removed due to low EMG/Acc data quality.

### 3.2 Activation of motor areas

The standard task regressor (a boxcar function convolved with the hemodynamic response function) and the EMG signal captured widespread activation within the sensorimotor system. Group-level analysis revealed motor task related BOLD activation across widespread cortical and subcortical regions, including the contralateral sensorimotor cortex (SMC), for the standard model m1 (Fig. 4).

**Fig. 4.**
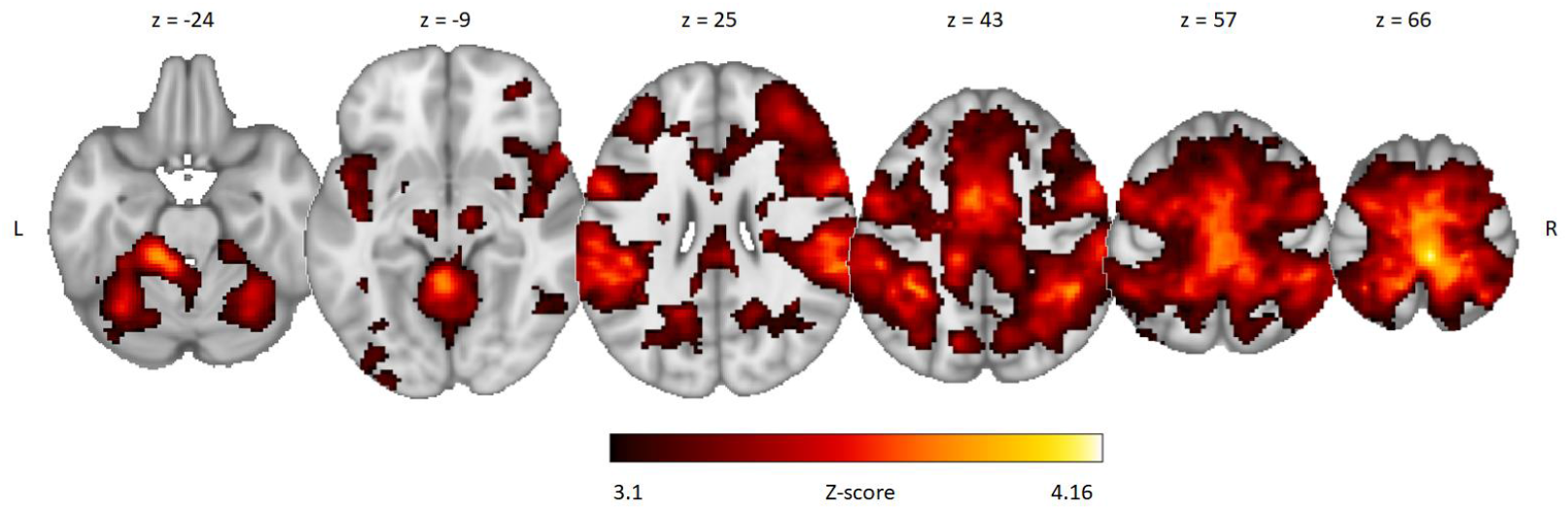
Task-related activations during ankle movement. Ankle condition as evaluated using model m1 with temporal derivative (n = 20; cluster-forming threshold Z > 3.1, cluster-wise p = 0.05 [family-wise error corrected], pre-threshold masking with standard brain mask) on top of the standard template. Please note that activations in models m1-m5 were essentially identical, hence only m1 is displayed.

### 3.3 Effect of rEMG, rAcc and split EMG (hypothesis h1)

Analysis of Acc- and rEMG-related activation revealed comparable effects of the orthogonalized regressors rAcc, rEMG and trEMG across models m2, m3, and m5, predominantly located in the sensorimotor cortex (Fig. 5, Table 1). Model m3 (rEMG) revealed two additional clusters located in the cerebellum and the right S2. In model m5 (split EMG), an additional cluster in the right precentral gyrus (dorsal premotor cortex, PMd, see Table 1) was detected, but activation was found neither in the cerebellum nor in the S2.

**Table 1.**
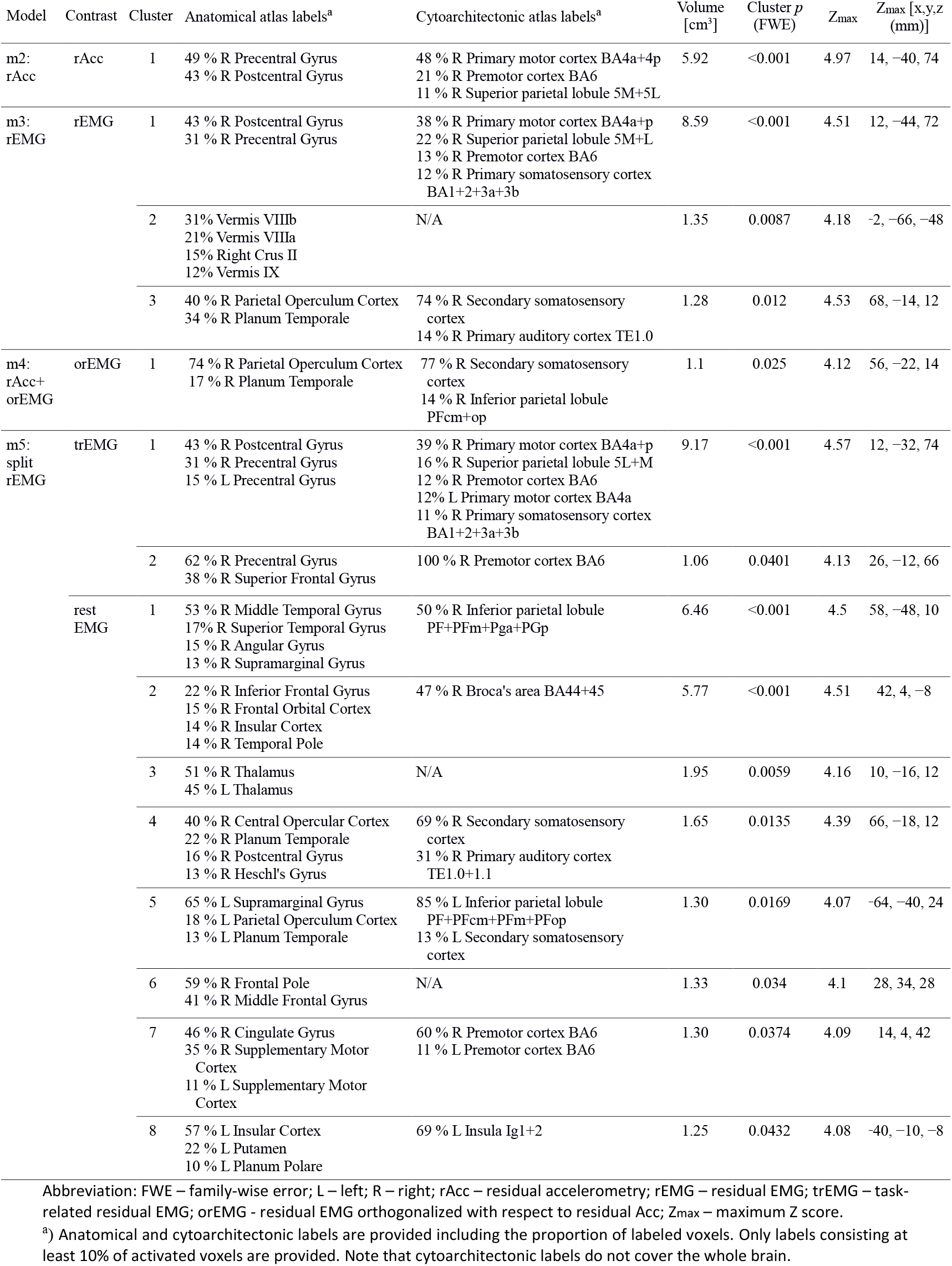
Group-wise effects in models – list of significant clusters and local maxima.

**Fig. 5.**
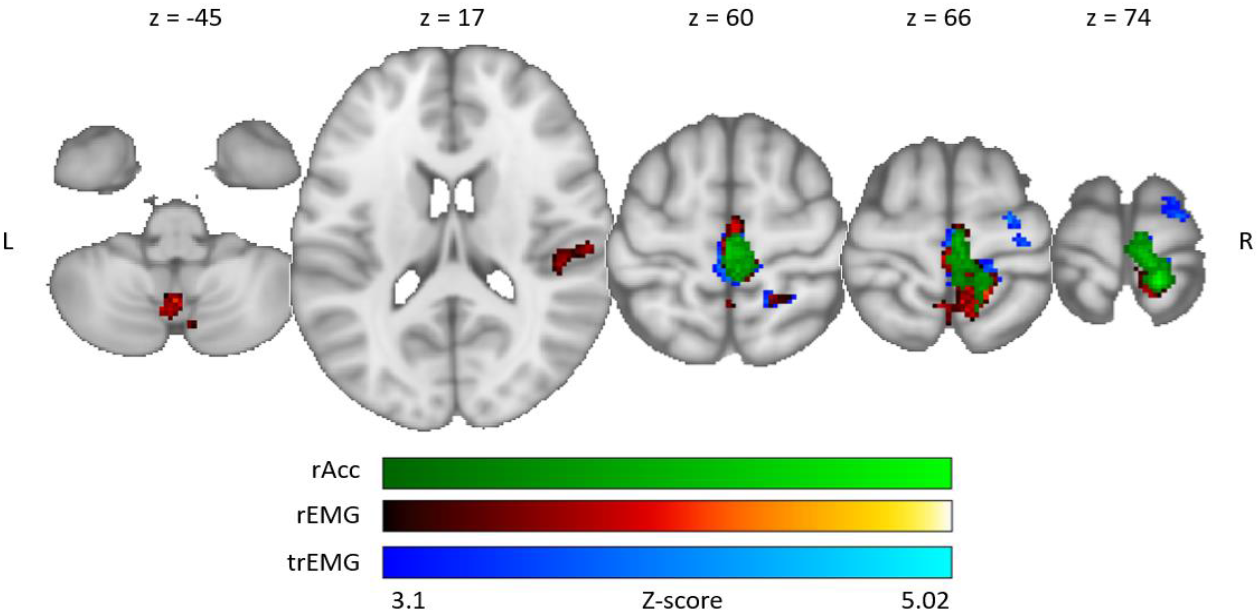
Acc- and EMG-related activations. Each overlay represents the main effect of the orthogonalized Acc or EMG regressor from a different model on top of the standard template: rAcc (m2; green), rEMG (m3; red), trEMG (m5; blue). All models include temporal derivatives (TD) of the main task regressor. Thresholded using cluster threshold Z > 3.1 (cluster-wise p = 0.05 [family-wise error corrected], pre-threshold masking with standard brain mask). Abbreviation: rAcc – residual accelerometry; rEMG – residual EMG; trEMG – task-related residual EMG.

Fig. 6 summarizes the post-hoc ROI analysis. In brief, significant non-zero activations were observed in bilateral SMC in all three comparable models (rAcc, rEMG, and split EMG). Model m3 (rEMG) exhibited additionally significant effects in the bilateral S2 (right S2: median [IQR] = 0.54 [0.77], p = 0.0041, r = 0.64; left S2: median [IQR] = 0.53 [1.08], p = 0.01, r = 0.58) and SMA (median [IQR] = 0.40 [0.98], p = 0.0251, r = 0.50), whereas model m5 (split EMG) showed only a borderline non-significant effect in the SMA (median [IQR] = 0.26 [0.92], p = 0.0569, r = 0.43).

**Fig. 6.**
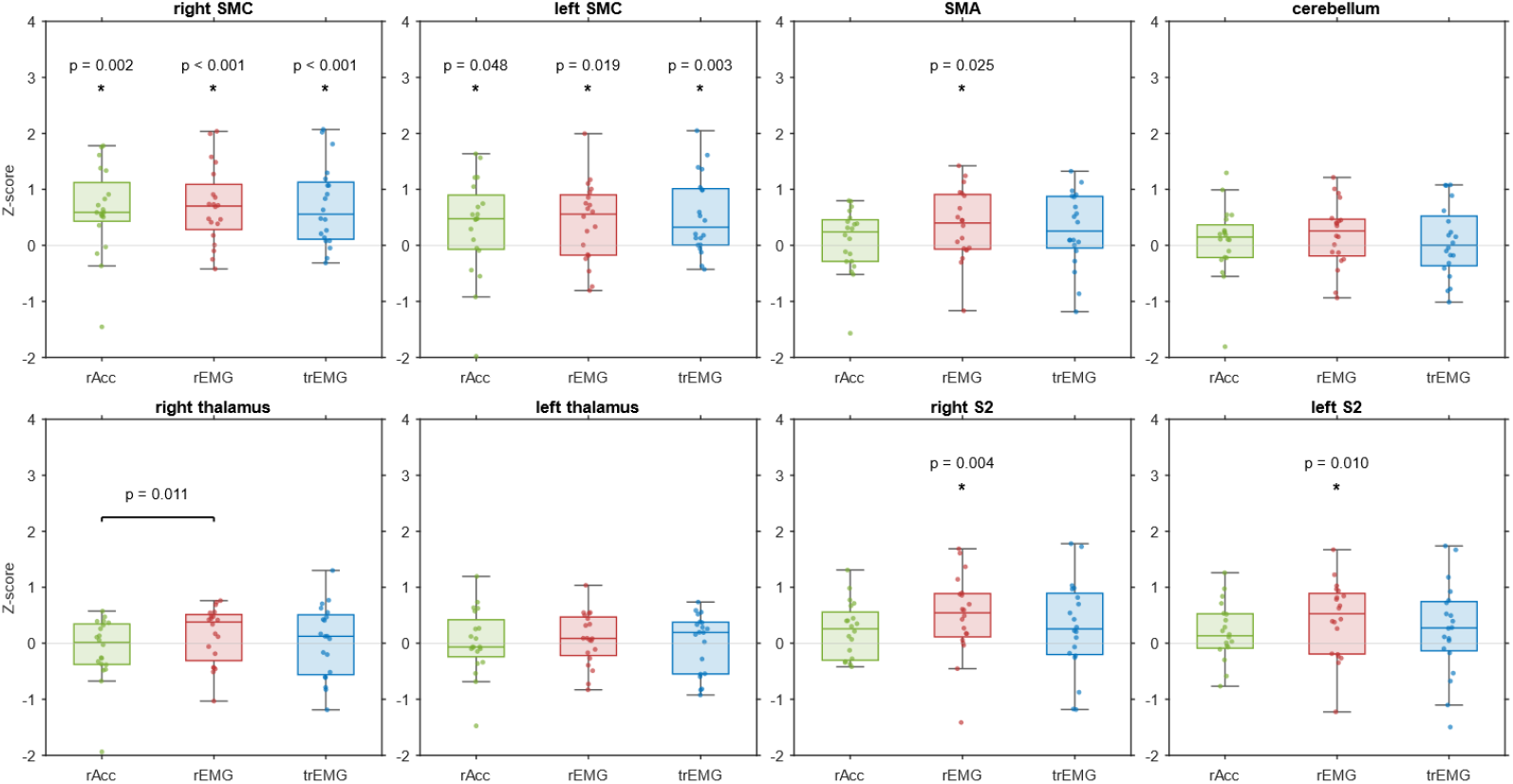
Statistical results. Boxplots show Z-scores for three analysis models across ROIs, each in a separate plot with a shared y-axis for direct comparison. Colors represent models m2 (rAcc; green), m3 (rEMG; red), m5 (split EMG; blue). All models included temporal derivatives of the main task regressor. Significant (p < 0.05) pairwise model differences are shown by horizontal lines with ticks and p-values; deviations from zero are marked by an asterisk with p-value. All statistics are based on non-parametric Wilcoxon signed-rank tests. Abbreviation: SMA – supplementary motor area; SMC – sensorimotor cortex; S2 – secondary somatosensory cortex; rAcc – residual accelerometry; rEMG – residual EMG; ROIs – regions of interest; trEMG – task-related rEMG.

### 3.4. Differences between models with rAcc, rEMG and split EMG (hypothesis h2)

Pairwise voxel-wise comparisons using FLAME 1+2 did not reveal any significant differences among the rAcc, rEMG and trEMG regressors (models m2, m3, and m5, respectively). In the ROI analysis, non-parametric Wilcoxon signed-rank test indicated a statistically significant difference between rAcc and rEMG in the right thalamus (p = 0.011, r = −0.57), as shown in Fig. 6. Two additional regions (in the bilateral S2) showed borderline effects but did not reach statistical significance (right S2: p = 0.0569, r = 0.43; left S2: p = 0.0522, r = 0.43).

### 3.5 Effect of orEMG (hypothesis h3)

Group-level analysis using FLAME 1+2 revealed one significant cluster (Fig. 7, Table 1) surviving cluster-wise correction for multiple comparisons, located in the right S2, with temporal derivatives included in the model. No additional significant clusters were observed. In the ROI analysis, a statistically significant non-zero activation was observed in the right S2 (median [IQR] = 0.52 [1.44], p = 0.0251, r = 0.5), right thalamus (median [IQR] = 0.28 [0.74], p = 0.0276, r = 0.49) and left S2 (median [IQR] = 0.63 [1.1], p = 0.0304, r = 0.48).

**Fig. 7.**
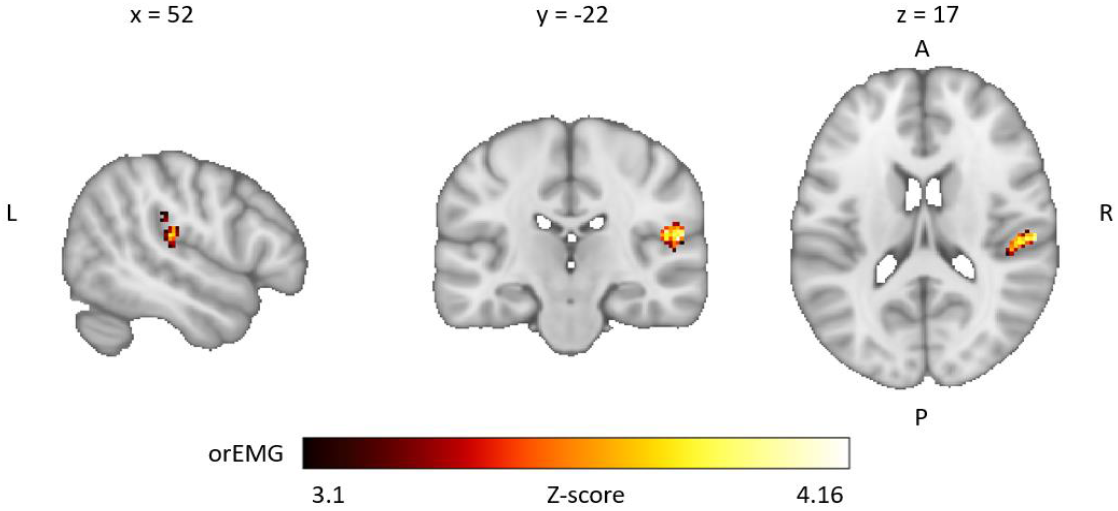
Effect of the orEMG in the model m4 (rAcc+orEMG). Statistical map showing activation explained by the rEMG in a model containing both rAcc and rEMG, with temporal derivative of the ankle movement task regressor (n = 20; cluster threshold Z > 3.1, cluster-wise p = 0.05 [family-wise error corrected], pre-threshold masking with standard brain mask). Abbreviation: Acc – accelerometry; orEMG – residual EMG orthogonalized with respect to residual Acc.

### 3.6 Effect of rest EMG (hypothesis h4)

Group-level analysis of effect of rest EMG in model m5 (split EMG) using FLAME 1+2 revealed eight significant clusters (Fig. 8, Table 1) at pFWE < 0.05, with temporal derivative of the task regressor included in the model. The largest clusters were located in the right temporoparietal junction and the right inferior frontal and insular cortex. Additional clusters were observed in the right S2, right prefrontal cortex, predominantly right SMA and cingulate cortex, left supramarginal gyrus, left posterior insular cortex, and bilateral thalami. In the ROI analysis, Wilcoxon signed-rank revealed a statistically significant non-zero activation in the cerebellum (median [IQR] = 0.28 [0.72], p = 0.0303, r = 0.48). A second region, right S2, showed a borderline effect (median [IQR] = 0.77 [1.73], p = 0.0569, r = 0.42) that did not reach statistical significance, while no significant effects were observed in the remaining ROIs.

**Fig. 8.**
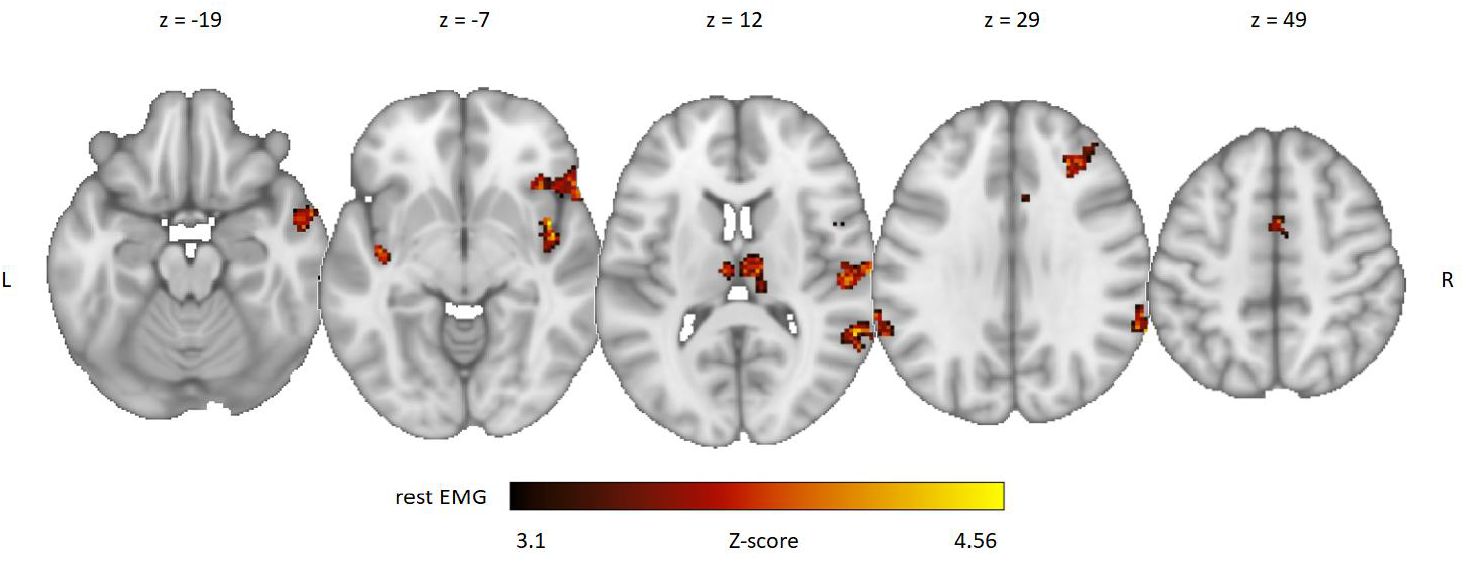
Effect of the rest EMG in model m5 (split EMG). Statistical map showing activation explained by the rest EMG regressor in the model separately modeling task-related rEMG and rest EMG, with temporal derivative of the ankle movement task regressor (n = 20; cluster threshold Z > 3.1, cluster-wise p = 0.05 [family-wise error corrected], pre-threshold masking with standard brain mask). Abbreviation: rEMG – residual EMG.

### 3.7 Effect of temporal derivatives of the main task regressor (hypothesis h5)

Wilcoxon signed-rank tests comparing effects of rAcc/rEMG regressors in the models with and without TD revealed statistically significant differences across several ROIs, as summarized in Table 2. Significant differences were observed in the cerebellum and in the right thalamus across tested models. Additionally, effects of the main task regressor were also compared across all models. Differences were observed in the right SMC and bilateral thalami.

**Table 2.** Effect of temporal derivatives – with TD vs. without TD across ROIs and models.

| ROI | physiological signal regressors |  |  |  |  | main task regressor |  |  |  |  |
| --- | --- | --- | --- | --- | --- | --- | --- | --- | --- | --- |
|  | m2<br>rAcc | m3<br>rEMG | m4<br>orEMG | m5<br>trEMG | m5<br>rest EMG | m1 | m2 | m3 | m4 | m5 |
| R SMC | 0.204<br>0.16±0.68 | 0.156<br>0.11±0.58 | 0.681<br>0.00±0.30 | 0.179<br>0.09±0.55 | 0.433<br>0.02±0.28 | **0.033**<br>0.13±0.21 | **0.025**<br>0.11±0.19 | **0.015**<br>0.10±0.23 | **0.021**<br>0.13±0.22 | 0.073<br>0.10±0.23 |
| L SMC | 0.263<br>0.13±0.77 | 0.218<br>0.06±0.59 | 0.765<br>-0.03±0.30 | 0.391<br>0.02±0.56 | 0.277<br>0.03±0.29 | 0.156<br>0.09±0.22 | 0.086<br>0.10±0.18 | 0.062<br>0.10±0.25 | **0.044**<br>0.11±0.21 | 0.135<br>0.10±0.26 |
| SMA | 0.654<br>0.05±0.45 | 0.351<br>0.06±0.32 | 0.391<br>0.30±0.23 | 0.478<br>0.04±0.30 | 0.550<br>0.01±0.18 | 0.478<br>0.02±0.19 | 0.263<br>0.05±0.17 | 0.355<br>0.04±0.13 | 0.204<br>0.05±0.15 | 0.314<br>0.05±0.12 |
| cerebellum | **0.005**<br>-0.21±0.51 | **0.011**<br>-0.21±0.40 | 0.108<br>-0.03±0.21 | **0.017**<br>-0.14±0.39 | **0.007**<br>-0.09±0.15 | 0.117<br>-0.04±0.20 | 0.296<br>-0.03±0.16 | 0.117<br>-0.03±0.19 | 0.156<br>-0.02±0.13 | 0.064<br>-0.03±0.16 |
| R S2 | 0.108<br>-0.14±0.61 | 0.240<br>-0.03±0.42 | 0.654<br>-0.05±0.18 | 0.279<br>-0.01±0.30 | 0.279<br>-0.01±0.16 | 0.601<br>-0.03±0.13 | 0.867<br>0.00±0.12 | 0.794<br>0.01±0.14 | 0.794<br>0.00±0.13 | 0.763<br>-0.01±0.15 |
| L S2 | 0.654<br>0.05±0.57 | 0.793<br>0.01±0.23 | 0.279<br>-0.05±0.15 | 0.681<br>-0.02±0.18 | 0.794<br>-0.01±0.07 | 0.433<br>0.03±0.15 | 0.422<br>0.02±0.13 | 0.218<br>0.03±0.15 | 0.433<br>0.03±0.15 | 0.279<br>0.02±0.18 |
| R thalamus | **0.021**<br>-0.19±0.35 | 0.135<br>-0.06±0.29 | 0.852<br>0.00±0.19 | 0.145<br>-0.06±0.27 | 0.126<br>-0.03±0.18 | **0.021**<br>-0.06±0.16 | **0.030**<br>-0.07±0.10 | **0.049**<br>-0.06±0.14 | **0.032**<br>-0.07±0.11 | **0.023**<br>-0.07±0.12 |
| L thalamus | 0.179<br>-0.05±0.40 | 0.263<br>-0.01±0.29 | 0.823<br>0.02±0.11 | 0.332<br>-0.02±0.23 | 0.167<br>-0.02±0.18 | 0.073<br>-0.03±0.21 | 0.135<br>-0.03±0.17 | 0.067<br>-0.03±0.18 | 0.079<br>-0.03±0.15 | **0.021**<br>-0.03±0.17 |

Abbreviation: L – left; R – right; rAcc – residual accelerometry; rEMG – residual EMG; ROIs – regions of interest; orEMG – rEMG orthogonalized with respect to rAcc; SMA – supplementary motor area; SMC – sensorimotor cortex; S2 – secondary somatosensory cortex; trEMG – task-related rEMG.
Statistically significant values ( $p < 0.05$ ), based on non-parametric Wilcoxon signed-rank tests, are highlighted in bold. In each cell, the first row shows the p-values, while the second row reports the median ± interquartile range (IQR) of the difference (with TD - without TD).

Despite these differences, most results of the main analysis with TD were still reproducible when TD of the main task regressor was left out from the models (see Supplementary Fig. S1-S4, showing data analogous to Fig. 5-8) with two exceptions: First, no rAcc-related activation was observed in the SMC. Second, rAcc, rEMG, orEMG, and trEMG yielded additional significant clusters in the cerebellum and ipsilateral hemisphere (see Supplementary Fig. S2-S4).

## 4. DISCUSSION

Motor task performance has infrequently been addressed in EMG- and kinematic-fMRI studies. Only a limited number of studies have explicitly modeled EMG variability using physiological signal regressors (van Rootselaar et al., 2007), whereas most previous work has treated EMG and kinematic measures as main task regressors reflecting mean motor output (e.g., MacIntosh et al., 2004) or as an off-line control parameter or covariate (e.g., Lotze et. al, 1999). Notably, van Rootselaar et al. (2007) demonstrated that rEMG variability represents a functionally meaningful signal rather than noise. The majority of previous EMG-fMRI and kinematic-fMRI studies have focused on upper limb movements, while to our knowledge, none studies have jointly modeled EMG- and kinematic-derived during the same movement.

In this study, we have assessed the effect of inclusion of physiological signals (rAcc and/or rEMG) into a model of BOLD signal changes during a simple foot movement. Main results include: (1) Orthogonalized (i.e., residual) Acc and EMG (rAcc and rEMG, respectively) from TA consistently explained BOLD signal changes in the mainly contralateral SMC (Fig. 5). (2) Signal changes explained by rAcc and rEMG do not differ on the whole brain level, although minor differences can be observed in the (contralateral) right thalamus (rEMG > rAcc; Fig. 6). rEMG, however, explained signal changes in several additional areas that were not detected in the model with rAcc, namely in the cerebellum, S2, and SMA (Fig. 5 and 6). (3) When rEMG was included on top of the rAcc within a single model as an orEMG regressor, it explained signal changes in the right S2 (Fig. 7). (4) BOLD signal changes in bilateral S2 and thalami were, however, better explained by the rest EMG in a model distinguishing between the task-related and resting EMG (Fig. 8). (5) Finally, several (but not all) results were dependent on the presence of the TD of the main task regressor (Table 2). Without TD, analysis was less sensitive to task-related activation in the predominantly contralateral SMC and thalamus. Additionally, rAcc and rEMG explained signal in multiple cerebellar regions and several additional areas, including bilateral temporal and occipital cortices and anterior cingulate gyrus, but signal changes explained by the rEMG in the SMC were less prominent or even absent, as in the case of rAcc.

### 4.1 BOLD signal changes explained by rAcc and rEMG in the SMC

Although we observed robust sensorimotor task-related activations irrespective of the physiological signals, the detected EMG-/Acc-related activations indicate that SMC additionally reflects moment-to-moment variations in muscle activity and the resulting movement execution (Fig. 5). This is in line with previous evidence that M1 preferentially encodes low-level movement features, such as movement amplitude (Stark-Inbar & Dayan, 2017), muscle activation level (Liu et al., 2000; MacIntosh et al., 2007). While with current analysis, it is not possible to determine whether these BOLD signal variations originate from endogenous fluctuations within the cortical loops or are driven by sensory feedback, they confirm that both Acc and EMG based on signal amplitude envelope convey information closely related to neuronal activity in the primary motor cortex (Liu et al., 2000; van Rootselaar et al., 2007).

While this finding may seem trivial, our novel approach with simultaneous Acc and EMG revealed that these physiological covariates result in strikingly similar recruitment of the SMC and that they do not differ on the whole brain level (MacIntosh et al., 2004), such that they might be considered as interchangeable in some applications in healthy subjects, e.g., for monitoring simple phasic movements along a single axis.

One consideration is that rAcc and rEMG could mainly reflect delay of the movement initiation. If that were the case, inclusion of the TD of the main task regressor should decrease the amount of BOLD signal variance explained by the rAcc or rEMG. The fact that we observed the opposite, suggests indeed that both rEMG and rAcc reflect chiefly fluctuations during the task block. Of note, effects in the SMC related to the rAcc but not those explained by the rEMG disappear completely when TD of the main regressor is removed, meaning that rEMG might be a more robust option less sensitive to some model settings. To our knowledge, temporal derivatives of physiological signals have not been explicitly modeled in previous EMG–fMRI or kinematic–fMRI studies and therefore represent a novel finding.

### 4.2 Differences between rAcc and rEMG outside the SMC

Despite the striking similarities between the rAcc- and rEMG-related activations in the SMC and the lack of significant voxel-wise differences on the whole-brain level, several complementary findings indicated that rEMG explained additional BOLD signal variability outside the primary sensorimotor areas (Fig. 5-7). First of all, adding rEMG on top of the rAcc as a twice orthogonalized regressor (orEMG) revealed a significant effect in the right (contralateral) S2 (Fig. 7), which was also supported by a borderline difference between rEMG and rAcc in the ROI analysis (Fig. 6). The left S2 showed in general similar effects on the ROI level, but it did not reach significance on the whole-brain level. Additionally, the ROI analysis identified an increased activation in the right thalamus for rEMG compared to rAcc (Fig. 6), which was confirmed by a significant effect for orEMG. Lastly, clusters in the cerebellar vermis VIII and in the right dorsal premotor cortex (Fig. 5) and an effect in the SMA on the ROI-level (Fig. 6) were detected for rEMG or trEMG, but not for rAcc.

In general, simple ankle movements engage (on top of SMC) the premotor cortices and the supplementary motor area for movement planning, coordination, and temporal control, the cerebellum for timing and fine modulation of muscle activity, and posterior parietal regions for proprioceptive and spatial integration (Liu et al., 2000; Ciccarelli et al., 2005). In addition, S2 and the thalamus contribute to the (higher-order) processing and relay of sensory and internal motor-related signals associated with ankle movement (Dobkin et al., 2004; Ciccarelli et al., 2005; Grooms et al., 2019).

In this study, the EMG-related activations in the contralateral S2 and thalamus coincided with similar activation reflecting spontaneous EMG fluctuations at rest (Fig. 5 and Fig. 8), which may explain the apparent discrepancy between rAcc and rEMG in these regions. Since Acc inherently reflects only overt limb movements, it cannot capture neuronal variability in the absence of measurable motion. In contrast, EMG-derived signals retain sensitivity to central motor processes and afferent feedback even during minimal or absent movement, as demonstrated in studies using isometric or low-motion paradigms (Liu et al., 2000; Ciccarelli et al., 2005).

Activations in the S2 and thalamus have been reported not only during active ankle movements but also under passive or minimal movement conditions (Ciccarelli et al., 2005; Dobkin et al., 2004) and can be observed during somatosensory foot stimulation (Hok et al. 2019). The engagement of S2 and thalamus during both active task and rest therefore suggests a contribution to sensorimotor integration or processing of sensory feedback (i.e., reflecting sensory consequences of motor output) rather than direct motor execution (Ciccarelli et al., 2005; MacIntosh et al., 2007).

The remaining regions observed for rEMG (cerebellum and premotor cortices, including the SMA, Fig. 5) were not associated with rest EMG. In fact, the premotor cortex was only observed for trEMG, suggesting its closer involvement in movement execution. However, the lack of significant differences between rEMG and rAcc in the premotor regions and cerebellum prevents us from drawing any definite conclusions. It thus remains to be seen whether these regions convey any specific information associated with EMG.

### 4.3. BOLD signal associated with rest EMG

Rest periods were defined as intervals outside the ankle movement condition, including a 3 seconds margin before and after movement condition. Given the experimental instructions, these periods may have included motor gait imagery in addition to two explicit rest conditions, and should therefore be interpreted as periods without overt movement rather than as an absence of motor-related neural activity (Guillot et al., 2012).

The observed activation pattern, including right S2, right prefrontal cortex, predominantly right SMA and cingulate cortex, left supramarginal gyrus, left posterior insular cortex, and bilateral thalami (Table 1), closely overlaps with networks previously reported during gait imagery and internally guided locomotor processing. Functional imaging studies have shown that gait imagery engages premotor and SMA, cingulate cortex, S2 and parietal regions, insula, and thalamus, even in the absence of overt movement (van der Meulen et al., 2014; Sacheli et al., 2020). Our data thus indicate that resting EMG might potentially capture sub-threshold motor activity associated with motor imagery, warranting further investigation.

### 4.3. Effects of TD in the model

TD of the main regressors were included to account for small temporal shifts between modeled and measured responses, especially to model potential latency variations and to reduce amplitude bias due to timing discrepancies (Calhoun et al., 2004; Pernet, 2014). In line with this, task-related activation in the SMC increased with TD without any significant impact on rEMG-related activation, suggesting that TD mainly improved model fit by explaining residual noise. Nevertheless, our data indicate that BOLD signals from several regions contain information of potentially neuronal origin that was removed by the TD regressor. Namely, inclusion of TD led to decreased activation in the cerebellum and thalami (Table 2). Whereas in thalami, TD captured signal variability in the main task regressor, in the cerebellum, TD of the task regressor explained a significant portion of the rAcc/rEMG-related variability. Hence, the BOLD responses in the cerebellum and the thalamus seem to involve brief signal changes reflecting transitions between the rest and movement (Li et al., 2021), with the cerebellum likely being more tightly coupled with the actual motor output.

Given the task-related activation increase in the SMC with TD, the results underscore the potential benefits of accounting for temporal variability in GLM-based fMRI analyses, particularly in studies involving clinical populations or naturalistic tasks, where timing variability may be more pronounced. Studies have shown that TD terms not only reduce residual variance due to slice-timing or motion (Soares et al., 2022), but also help eliminate amplitude bias when there is misalignment between the model and the actual hemodynamic response (Calhoun et al., 2004). At the same time, however, TD may potentially capture and thus conceal the true neuronal signal with more transient characteristics, as in the case of the cerebellum or thalami in our study.

### 4.4 Implications for studies in patient populations

The results highlight the potential of the current methods to quantify and isolate distinct neural correlates, supporting the use of combined fMRI and EMG as a promising approach for developing new biomarkers of both central and peripheral motor control impairments. By explaining a significant portion of the BOLD signal variance, the EMG regressor provides a link between peripheral motor output and central neural processes, bridging the gap between brain activation and actual movement execution. This work serves as a basis for studies focusing on the highly topical issues of low interpretability and limited reliability of fMRI, as well as the ongoing need for objective monitoring of biomarkers in neurological diseases (e.g., multiple sclerosis, stroke). The scope of the current study allows for a detailed initial evaluation of the EMG-fMRI methodology for the diagnosis of lower limb paresis and gait disorders and will form the basis for further clinical studies monitoring the effect of individual interventions.

### 4.5 Limitations

A limitation of this study is the exclusive focus on healthy controls, which understandably led to expected patterns of activation in motor-related brain areas. While this outcome supports the validity of the proposed methods, the findings primarily serve as a foundation for future investigations involving patients with gait disorders, where more clinically meaningful insights are anticipated. An a priori power analysis was not conducted, and thus the absence of statistically significant effects cannot be interpreted as evidence of their true absence. The study was conceived as an exploratory, pilot investigation aimed at testing the feasibility of the experimental design and analysis framework. Accordingly, the effect sizes observed here may be used to guide power estimations and sample size planning in subsequent confirmatory studies.

In addition, EMG recordings were limited to a single muscle (TA), and the analysis relied on a specific preprocessing pipeline; recordings from additional muscles and alternative EMG processing strategies may provide complementary insights and should be explored in future studies.

## 5. Conclusions

EMG-fMRI recordings have drawn growing interest in clinical and biomechanical research. The results of this study highlight the potential of this multimodal approach to deepen our understanding of the physiology of motor control, particularly in relation to lower limb mobility. The findings demonstrate the viability of developing robust and potentially clinically relevant metrics for detection and evaluation of movement impairments. Furthermore, we propose several novel approaches for incorporating EMG into the GLM analysis of BOLD MRI signals to increase interpretability, such as separate modeling of the task-related and resting EMG or double orthogonalized EMG. This represents a meaningful advancement in studying both normal motor system function and the pathophysiological mechanisms underlying movement disorders. Taken together, these findings demonstrate that incorporating EMG and Acc signals into fMRI analysis enables identification of brain regions whose activity is tightly coupled to peripheral motor output. This integrative approach provides a more nuanced view of motor control, linking central neural activity with real movement execution, and holds substantial potential for future pathophysiological mechanisms.

## Data Availability

All data produced in the present study are available upon reasonable request to the authors.

## Acknowledgments

We acknowledge the core facility MAFIL supported by MEYS CR (LM2023050 Czech-BioImaging), part of the Euro-BioImaging (www.eurobioimaging.eu) ALM and Medical Imaging Node (Brno, CZ). MJ, PeH and BK were supported by MH CZ – DRO (FNOL, 00098892). PaH was supported by the Grant Agency of Masaryk University project MUNI/SC/1939/2024. AH was supported by the EU HE HybridNeuro project (101079392) and by Slovenian Research and Innovation Agency (J2-60046, P2-0041). The authors would like to thank Kristína Ruszová, Klára Balážová and Jana Bahrová for their helpful contributions throughout the project.

