## Supplementary material for "Precise modeling of task-related sensorimotor activation based on simultaneous surface electromyography"

### SUPPLEMENTARY FIGURES

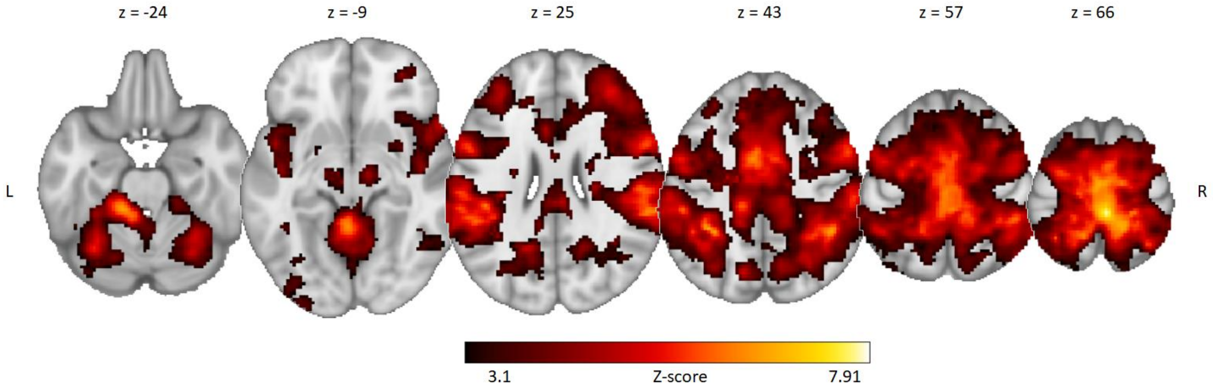


**Supplementary Fig. S1 Task-related activations during ankle movement.** Ankle condition as evaluated using model m1' without temporal derivative (n = 20; cluster-forming threshold Z > 3.1, cluster-wise p = 0.05 [family-wise error corrected], pre-threshold masking with standard brain mask) on top of the standard template. Please note that activations in models m1'-m5' were essentially identical, hence only m1' is displayed.

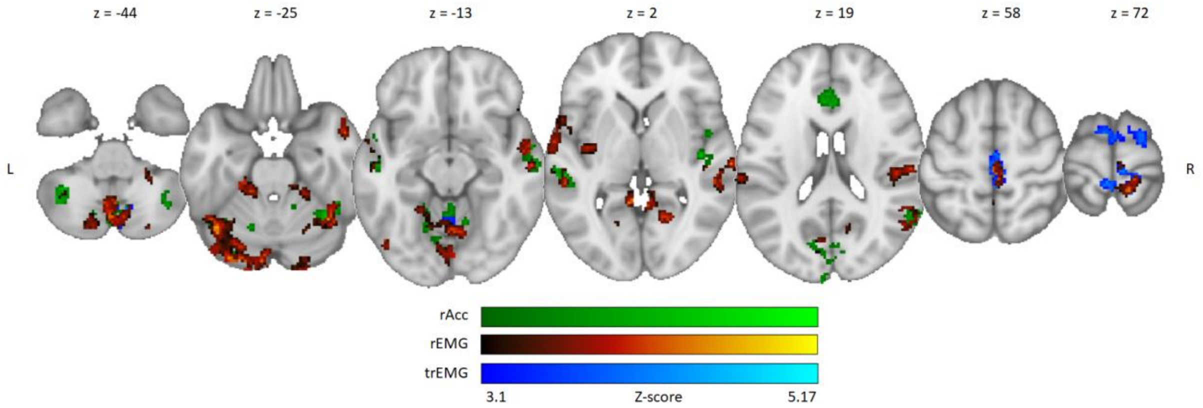


**Supplementary Fig. S2 Acc- and EMG-related activations.** n = 20; cluster threshold Z > 3.1, Zmax = 5.17, cluster-wise p = 0.05 [family-wise error corrected], pre-threshold masking with standard brain mask on top of the standard template. Each represents a different model: m2' (rAcc; green); m3' (rEMG; red), m5' (trEMG; blue). All included models are without temporal derivatives (TD) of the main task regressor. Abbreviation: rAcc – residual accelerometry; rEMG – residual EMG; trEMG – task-related residual EMG.

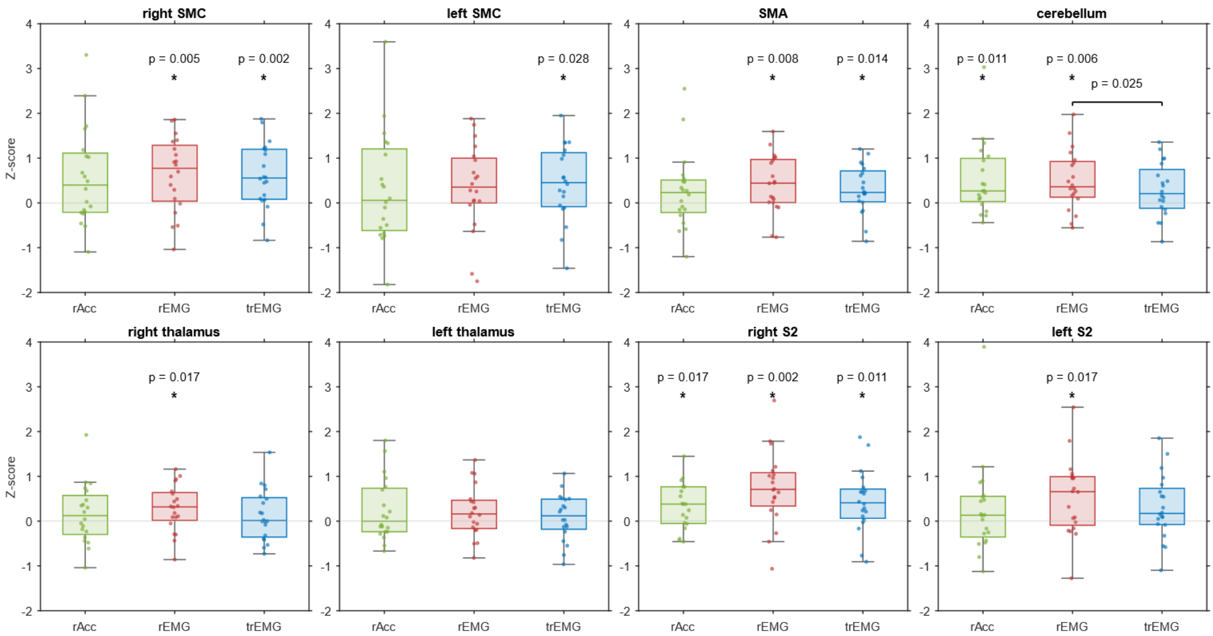


**Supplementary Fig. S3 Statistical results.** Boxplots show Z-scores for three analysis models across ROIs, each in a separate plot with a shared y-axis for direct comparison. Colors represent models m2' (rAcc; green), m3' (rEMG; red), m5' (split EMG; blue). All included models are without temporal derivatives of the main task regressor. Significant ($p < 0.05$) pairwise model differences are shown by horizontal lines with ticks and p-values; deviations from zero are marked by an asterisk with p-value. All statistics are based on non-parametric Wilcoxon signed-rank tests. Abbreviation: SMA – supplementary motor area; SMC – sensorimotor cortex; S2 – secondary somatosensory cortex; rAcc – residual accelerometry; rEMG – residual EMG; ROIs – regions of interest; trEMG – task-related rEMG.

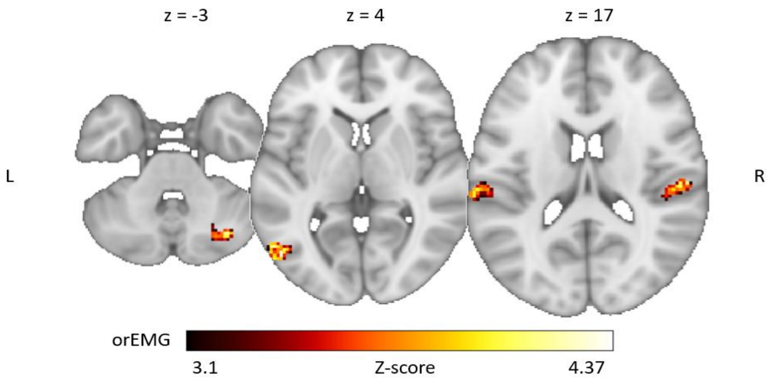


**Supplementary Fig. S4 Effect of orEMG in the model rAcc+orEMG (m5).** Statistical map showing activation explained by rEMG in a model containing both rAcc and rEMG, without temporal derivative of the ankle movement task regressor (n = 20; cluster threshold Z > 3.1, cluster-wise p = 0.05 [family-wise error corrected], pre-threshold masking with standard brain mask). Abbreviation: rAcc = residual accelerometer (time-series), rEMG = residual EMG; orEMG – residual EMG orthogonalized with respect to residual Acc.

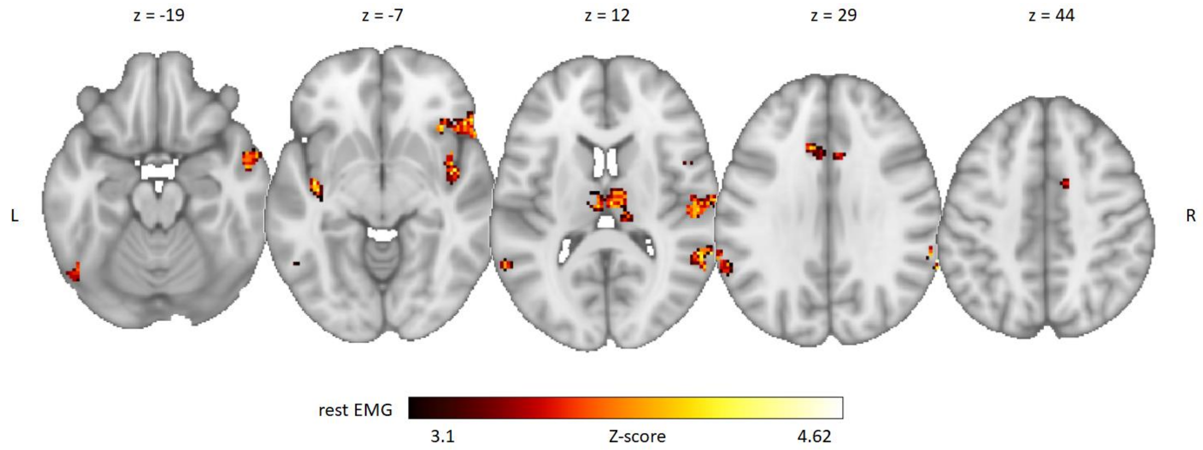


**Supplementary Fig. S5 Effect of rest EMG in model m4 (split rEMG).** Statistical map showing activation explained by rest EMG in a model separately modeling task-related rEMG and rest EMG, without temporal derivative of the ankle movement task regressor (n = 20; cluster threshold Z > 3.1, cluster-wise p = 0.05 [family-wise error corrected], pre-threshold masking with standard brain mask). Abbreviation: rEMG = residual EMG.
